# Diagnostic Accuracy of Preoperative C-reactive protein/albumin ratio for Predicting Postoperative Delirium: A Protocol for a Systematic Review and Diagnostic Test Accuracy Meta-analysis

**DOI:** 10.1101/2025.10.29.25339103

**Authors:** Takahiro Mihara, Makiko Okazaki, Ryo Koto, Hisashi Noma, Kanako Sasaki

## Abstract

**Background:** Postoperative delirium (POD) is a frequent and severe complication associated with increased morbidity, mortality, and healthcare costs. Effective preoperative risk stratification is crucial for prevention. The C-reactive protein/albumin ratio (CAR), an accessible biomarker that reflects both systemic inflammation and nutritional status, shows promise for POD prediction. However, the overall diagnostic performance across diverse surgical populations has not yet been quantified. This study aims to systematically review and synthesize evidence on the diagnostic accuracy of preoperative CAR in predicting POD.

**Methods:** This protocol will adhere to the PRISMA-DTA statement. A comprehensive search will be conducted in major electronic databases, including MEDLINE, Embase, and Cochrane Library, without language restrictions. Observational studies and clinical trials evaluating the accuracy of preoperative CAR in predicting POD in adult surgical patients, where POD is diagnosed using a validated tool, will be included. Two reviewers will independently perform study screening, data extraction, and risk of bias assessment using the QUADAS-2 tool. For the primary analysis, a bivariate random-effects model will be used to synthesize true-positive, false-positive, true-negative, and false-negative data, generating coupled forest plots and a hierarchical summary receiver operating characteristic (sROC) curve. Heterogeneity will be quantified, and its sources will be explored through pre-specified subgroup and sensitivity analyses.

**Ethics and Dissemination:** As a systematic review of previously published data, formal ethical approval is not required. The findings will be disseminated through publication in a peer-reviewed journal and presentation at scientific conferences.

## Introduction

Postoperative delirium (POD) is one of the most crucial and challenging complications in the perioperative period, imposing a substantial burden on patients, healthcare systems, and society [1,2]. Defined as an acute and fluctuating disturbance in attention, awareness, and cognition, POD is not merely a transient state of confusion but a serious medical condition linked to a cascade of devastating adverse outcomes [1–3]. Patients who develop POD face an increased risk of prolonged hospital stays, a greater need for institutional care after discharge, persistent long-term cognitive impairment, and a substantial increase in both short-term and long-term mortality [1–3]. The incidence of POD is alarmingly high and varies with the patient population and the nature of the surgical procedure. In high-risk cohorts, such as elderly patients undergoing major cardiac surgery or emergency orthopedic procedures for hip fractures, the incidence can exceed 50% [2,4,5]. This clinical reality underscores the urgent need for effective preventative strategies, which must be predicated on the accurate and early identification of at-risk individuals before surgery.

The pathophysiology of POD is known to be multifactorial, arising from a complex interplay of predisposing patient vulnerabilities and precipitating perioperative insults [2,6]. Among the leading hypotheses, the neuroinflammation model has gained considerable traction, supported by a growing body of clinical and preclinical evidence [7,8]. This model suggests that physiological stress during surgery triggers a potent systemic inflammatory response. Peripheral inflammation can disrupt the integrity of the blood-brain barrier, allowing inflammatory mediators to enter the central nervous system, thereby activating microglia and promoting a state of neuroinflammation that manifests clinically as delirium. This framework provided a strong rationale for investigating inflammatory biomarkers as potential predictors of POD risk.

The C-reactive protein/albumin ratio (CAR) has emerged as a promising candidate biomarker in this context [2,9]. The strength lies in its composition as a dual-domain indicator. C-reactive protein (CRP) is a highly sensitive, albeit non-specific, marker of acute systemic inflammation [10] that directly reflects the magnitude of the surgical inflammatory insult. Conversely, serum albumin serves as a robust indicator of the patient’s underlying physiological state, reflecting not only long-term nutritional status but also acting as a negative acute-phase reactant and surrogate for physiological reserve and frailty [11–14]. By integrating these two measures into a single ratio, CAR offers a more holistic assessment than either marker alone [15,16]. It theoretically captures both the patient’s baseline vulnerability (low albumin) and the intensity of the acute precipitating stressor (high CRP), elegantly mirroring the “two-hit” model of delirium pathogenesis.

A growing number of primary studies have explored the predictive utility of preoperative CAR for POD across various surgical disciplines, including orthopedics [17] and general surgery[9]. These individual studies have often reported that an elevated preoperative CAR is an independent predictor of POD. However, this body of evidence remains fragmented and is limited. Previous studies were single-center, retrospective in design, and confined to specific, often homogeneous surgical populations. Consequently, the overall diagnostic performance of CAR remains poorly defined and its generalizability across the broad and heterogeneous landscape of surgical practice is unknown. To date, no systematic review or meta-analysis has been conducted to synthesize the disparate findings. This represents a critical knowledge gap, hindering the successful implementation of CAR into routine preoperative risk assessment pathways.

Therefore, this protocol outlines the methodology for a comprehensive systematic review and diagnostic test accuracy (DTA) meta-analysis to formally evaluate the predictive performance of preoperative CAR for POD in adult surgical patients. The primary aim is to ascertain overall diagnostic accuracy by deriving pooled estimates of sensitivity and specificity and constructing a summary receiver-operating characteristic (sROC) curve to calculate the area under the curve (AUC) as a comprehensive measure of test performance. Additionally, we will investigate the sources of heterogeneity through pre-specified subgroup analyses. By providing a robust, consolidated estimate of its diagnostic properties, this review aims to clarify the clinical value of CAR as a simple, inexpensive, and universally available tool for preoperative risk stratification, with the goal of facilitating targeted interventions to prevent this devastating complication.

## Methods

This manuscript was prepared according to the recommendations of the Preferred Reporting Items for Systematic Review and Meta-analysis of Diagnostic Test Accuracy Studies (PRISMA-DTA) [18]. The protocol for this systematic review was registered with PROSPERO (registration number CRD420251171160) on October 19, 2025. Ethical approval was not required as this systematic review used previously published data.

### Eligibility criteria

#### Study design

We will include retrospective and prospective observational studies as well as randomized controlled trials, without restricting our search to specific languages. Case reports, case series, and animal studies will be excluded.

#### Patients

We will include studies on adult patients (≥18 years) who underwent any type of surgery.

#### Index tests

The index test is the C-reactive protein/albumin ratio (CAR). CAR is a biomarker derived from routine blood tests, calculated as the serum C-reactive protein (CRP) level divided by the serum albumin level. It is considered an indicator of the systemic inflammatory response and nutritional status of the patient.

#### Target condition

The target condition is postoperative delirium (POD). The diagnosis of POD will be based on the definition used in each primary study, provided that a validated screening or diagnostic tool (e.g., Confusion Assessment Method [CAM], CAM-ICU) was used.

### Information sources and search strategy

We will search MEDLINE, Embase, the Cochrane Central Register of Controlled Trials (CENTRAL), ClinicalTrials.gov, the European Union Clinical Trials Register, the WHO International Clinical Trials Registry Platform, and the UMIN database for appropriate studies. Moreover, we will manually search the reference lists of the relevant articles. The final search will be conducted on October 22, 2025.

The search strategy will involve a combination of free-text words and Medical Subject Headings (MeSH) terms across two main concepts. The first concept will include terms for the index test components, such as “c reactive protein” and “serum albumin”. The second concept will encompass a broad range of terms for the target condition, including “Delirium,” “postoperative delirium,” “acute confusional state” and validated assessment tools like “Confusion Assessment Method.” Methodological search filters will be avoided to maximize sensitivity, and no language restrictions will be applied. In this protocol, we present the PubMed (MEDLINE) search strategy in Table 1. Strategies for other databases will be submitted as supplementary materials for journal submission.

**Table 1:** The search strategy for PubMed (MEDLINE)

| # | Concept | Search terms (PubMed fields) |
| --- | --- | --- |
| #1 | Biomarker | ((("c reactive protein"[MeSH Terms] OR "c reactive protein"[Title/Abstract] OR "CRP"[Title/Abstract]) AND ("serum albumin"[MeSH Terms] OR "albumin"[Title/Abstract] )) |
| #2 | Delirium | ("Delirium"[MeSH Terms] OR "Delirium"[Title/Abstract] OR "postoperative delirium"[Title/Abstract] OR "post-operative delirium"[Title/Abstract] OR "POD"[Title/Abstract] OR "acute confusional state*"[Title/Abstract] OR |
|  |  | "acute confusion"[Title/Abstract] OR "hypoactive delirium"[Title/Abstract] OR "hyperactive delirium"[Title/Abstract] OR "mixed delirium"[Title/Abstract] OR "Confusion Assessment Method"[Title/Abstract] OR "CAM"[Title/Abstract] OR "CAM-ICU"[Title/Abstract] OR "ICDSC"[Title/Abstract] OR "Intensive Care Delirium Screening Checklist"[Title/Abstract] OR "Nursing Delirium Screening Scale"[Title/Abstract]) |
| #3 | Search strategy | #1 AND #2 |

### Study records

#### Data management and selection process

Search results from various databases will be compiled in Rayyan [19], where duplicates will be identified and removed. At each stage (title/abstract and full-text screening), two of the three authors (TM, MO, and RK) independently screened records against the eligibility criteria. When study eligibility cannot be determined based on the title or abstract, the full text will be retrieved.

#### Data collection

Two authors (TM and KS) will extract the data independently and in duplicates from each eligible study. Any disagreements regarding eligibility or extracted data will be resolved through discussion. When necessary, we would contact the study authors to obtain detailed data if the studies reported only an association between CAR and POD, without sufficient data to construct a 2 × 2 contingency table. After obtaining the details, we will determine the counts of true-positives, true-negatives, false-positives, and false-negatives based on the threshold value reported in the primary study, and subsequently incorporate them into our meta-analysis. If a study reported multiple thresholds, we would adopt the one identified as optimal by the original authors.

#### Data items

A data collection sheet will be created to record the following information: author information, year of publication, patient recruitment period, study design, inclusion criteria, exclusion criteria, patient characteristics (age, sex, height, weight, and body mass index), type of surgery, type of anesthesia, postoperative destination categorized as ICU vs. non-ICU, with the proportion in each category, timing of CAR measurement, timing of POD diagnosis, cut-off value of the index test, definition and diagnostic tool for POD, the total number of patients, and the total number of true-positives, false-positives, false-negatives, and true-negatives for the given cut-off.

### Outcomes and prioritization

#### Primary outcome

The primary outcome is the diagnostic accuracy of preoperative CAR in predicting postoperative delirium. We will report summary predictive measures, such as sensitivity, specificity, and summary receiver operating characteristic (sROC) curves.

### Risk-of-bias in individual studies

The risk-of-bias will be assessed using the Quality Assessment of Diagnostic Accuracy Studies (QUADAS-2) tool [20]. The QUADAS-2 has four domains: patient selection, index test, reference standard, and flow and timing. Two reviewers (TM and KS) will independently assess the risk-of-bias of the studies included. Disagreements between them will be resolved through discussion.

### Data synthesis

First, we will summarize the outcome measures in individual studies based on the number of true-positives, false-positives, false-negatives, and true-negatives. We will calculate the sensitivities, specificities, and corresponding 95% confidence intervals (CIs). We will present coupled forest plots to depict sensitivities and specificities for CAR and to undertake initial exploratory evaluations and assess heterogeneities.

We will then perform synthesis analyses using Reitsma’s bivariate random effects [21] model for study-specific sensitivities and specificities to address possible heterogeneity across studies. Based on the results of the synthesis analyses, we will estimate the summary sensitivities and specificities and create sROC curves [22]. We will assess heterogeneity using the bivariate version of I^2^, considering the correlation between sensitivity and specificity [23]. We will present the areas under the curves (AUCs) of the sROC curves as summary measures of predictive accuracy. For statistical inferences, we will use the standard restricted maximum likelihood estimation for Reitsma’s model and the bootstrap method to calculate the 95% CIs of the AUCs of the sROC curves [24].

If heterogeneity exists, we plan to perform a subgroup analysis to explore the underlying causes. Subgroup analyses are designed according to the following variables: type of surgery (e.g., cardiac, orthopedic, abdominal), type of anesthesia, cut-off values for CAR, and the diagnostic tool used for POD. We will perform sensitivity analyses after excluding studies with a high risk-of-bias. To assess potential publication biases, we will perform a generalized Egger test for multivariate meta-analysis [25].

Statistical analyses will be performed using R software version 4.5.1 (R Development Core Team, Vienna, Austria) and RStudio (RStudio, Boston, Massachusetts, USA).

## Discussion

This systematic review will be the first to synthesize the diagnostic accuracy of preoperative CAR for predicting POD across various surgical populations. By providing pooled estimates of sensitivity and specificity, this study aims to clarify the clinical utility of CAR as a simple, low-cost, and widely available tool for preoperative risk stratification. These findings could inform clinical guidelines and promote a shift from reactive to proactive delirium management, allowing for the targeted allocation of preventive resources to high-risk patients. For example, proactive measures may include the use of dexmedetomidine [26]; postoperative non-pharmacologic bundles [27]; and melatonergic agents [28] (e.g., melatonin or ramelteon).

Potential limitations include expected heterogeneity across studies in terms of CAR cut-off values, surgical populations, and POD assessment methods. We will explore these factors through pre-planned subgroup analyses. Despite these challenges, this review will provide a comprehensive summary of the current evidence, identify key research gaps, and guide future studies in this essential clinical area.

## Data Availability

This protocol does not include any patient-level data; no datasets were generated or analyzed. Data sharing is therefore not applicable.

## Data and Code Availability

No original data were obtained.

## Author contributions

This study was designed by TM. TM was primarily responsible for writing the manuscript. All the authors critically revised the manuscript for important intellectual content and approved the final draft. All authors have read and approved the final version of the manuscript.

## Declaration of generative artificial intelligence (AI) in scientific writing

During the preparation of this manuscript, the authors used ChatGPT (OpenAI, San Francisco, CA, USA) to assist in improving the logical flow and overall structure of the text. After using this tool, the authors carefully reviewed and edited the content where necessary and took full responsibility for the final version of the manuscript.

